# Understanding Suicide in Our Community Through the Lens of the Pediatric ICU: An Epidemiological Review (2011-2017) of One Midwestern City in the US

**DOI:** 10.1101/2020.12.10.20247072

**Authors:** Andrew Kampfschulte, Matthew Oram, Alejandra M. Escobar Vasco, Brittany Essenmacher, Amy Herbig, Aniruddh Behere, Mara L. Leimanis-Laurens, Surender Rajasekaran

**Affiliations:** Office of Research and Education, Spectrum Health, 15 Michigan Street NE, Grand Rapids, MI, United States; Department of Pediatrics and Human Development, College of Human Medicine, Michigan State University, Life Sciences Building, 1355 Bogue Street, East Lansing, MI, 48824; Pediatric Intensive Care Unit, Helen DeVos Children’s Hospital, 100 Michigan Street NE, Grand 16 Rapids, MI, 49503; Pediatric Behavior Health, Helen DeVos Children’s Hospital, 100 Michigan Street NE, Grand 14 Rapids, MI, 49503

**Keywords:** suicide, pediatrics, critical care, geospatial, temporal, self-directed violence

## Abstract

Suicide frequency has tripled for some pediatric age groups over the last decade, of which, serious attempts result in pediatric intensive care unit (PICU) admissions. We paired clinical, aggregate geospatial, and temporal demographics to understand local community variables to determine if epidemiological patterns emerge that associate with risk for PICU admission. Data was extracted at an urban, high-volume, quaternary care facility from January 2011 to December 2017 via ICD 10 codes associated with suicide. Clinical, socioeconomic, geographical, and temporal variables were reviewed. 1,036 patients over age 9 were included, of which n=161 were PICU admissions. Females represented higher proportions of all suicide-related hospital admissions (67.9%). Looking at race/ethnicity, PICU admissions were largely Caucasian (83.2%); Blacks and Hispanics had lower odds of PICU admissions (OR: 0.49; 0.17, respectively). PICU-admitted patients were older (16.0 vs. 15.5; p=0.0001), with lower basal metabolic index (23.0 vs. 22.0; p=0.0013), and presented in summer months (OR: 1.51, p = 0.044). Time-series decomposition showed seasonal peaks in June and August. Local regions outside city limits identified higher numbers of PICU admissions. PICUs serve discrete geographical regions and are a source of information, when paired with clinical-geospatial/seasonal analyses, highlighting clinical and societal risk factors associated with PICU admissions.

## 1. Introduction

Suicide is defined by the Centers for Disease Control as death caused by injuring oneself with the intent to die and is part of a broader class of behaviors called self-directed violence, that is a behavior that could result in immediate injury and has potential for lasting injury [1]. A serious suicide attempt (SSA) is one that would result in death without specialized intervention (surgery, antidotes, intensive care unit (ICU) admission, prolonged hospitalization), and can be considered a proxy for a completed suicide in surviving individuals [2]. This further breakdown of patients into two groups may be summarized as: those requiring hospitalization for observation versus those facing potential injury or risk of mortality, thus requiring an ICU admission.

Suicide is a major and ongoing public health concern in the United States (US). Current data demonstrates that death by suicide is the second leading cause of mortality in people age 15-24 years [3,4]. Suicide affects young people from all races and socioeconomic groups. Rates of suicide have increased since the turn of the century in both adults and children [3]. From 1999-2014, the rate of suicide for children age 10-14 tripled, helping to drive the overall increase in rate of suicide across all ages during the same time period [5].

Children’s hospitals serve distinct geographical areas, where patients needing highly advanced care are sent to their regional ICU. We are the only pediatric ICU (PICU) in a city of above 200,000 residents with a catchment area that includes one million residents. Patients that originate from these regions are being exposed to a unique set of socioeconomic, geographical, and temporal factors that need better understanding and characterization.

In this retrospective chart review, we extracted clinical and community variables available in the electronic medical record (EMR) to develop a more complete understanding of factors that associate with PICU admission (non-ICU hospitalized patients were used as our comparative-control group). We hypothesized that since PICUs serve distinct regions, an analysis of the variables extracted from the EMR could assist in developing an understanding of clinical, socioeconomic, geographical [6,7], and temporal (yearly, seasonal, time of day, *-of incident*) [8], demographics for our region. The results presented herein are intended for PICU staff, pediatricians, social workers, chaplains, suicide researchers, parents, educators, psychologists, those working in adolescent community outreach, and mental health fields.

## 2. Materials and Methods

### 2.1 Site and Population

The study and data collection were conducted at Helen DeVos Children’s Hospital (HDVCH) located in Grand Rapids, MI. HDVCH PICU is a mixed cardiac surgery and medical intensive care unit with approximately 1600 annual admissions, with over 6,000 patient days. Seventeen board certified intensivists provide 24-hour coverage for a 24-bed unit, with maximum capability of 36 critically ill children.

Patients included in our study were between 9 and 18 years of age admitted to HDVCH between January 2011 to December 2017. Medical records were manually screened retrospectively for possible suicide-related diagnoses and admission keywords (overdose, poisoning, ingestion, intoxication, suicide attempt, or altered mental status). Relevant indications for ICU admission included respiratory failure, significant risk for respiratory failure, depressed mental status, seizures, cardiovascular dysfunction, and risk of arrhythmias. We excluded patients < 9 years and > 18 years of age, with developmental delays, and patients where intent of suicide was difficult to establish.

### 2.2 Variables

Data extracted from HDVCH’s EMR included categorical variables: suicide category (drowning, hanging/strangling, poison, drug overdose, gas, cutting, fall by height)], race of patients (African American, Caucasian, Hispanic, other), and type of insurance (unknown, commercial, government), discharge disposition (home, psychiatric/rehab, expired, other). Binary variables included sex (dummy variable: male =0; female=1), admission through the emergency department (ED) (Yes =1/ No=0), hospital death (Yes=1/No=0), death (Yes=1/No=0); numerical variables included basal metabolic index (BMI), age of the patient (years), and length of stay (LOS), median income (Supplemental Table 1). Severity of illness scores included pediatric risk of mortality III (PRISM III) score [9], and Pediatric Index of Mortality (PIM2) [10], both calculated during the first hours of ICU admission.

**Table 1:** Total Patient Demographics

| Variable | Levels | Missing N | No PICU | PICU | Total | *p* |
| --- | --- | --- | --- | --- | --- | --- |
| Total N (%) |  |  | 875 (84.5) | 161 (15.5) | 1036 |  |
| Age | Median (IQR) | 0 | 15.5 (2.6) | 16.0 (2.4) | 15.6 (2.5) | **0.001** |
| Gender | Female | 0 | 600 (68.6) | 103 (64.0) | 703 (67.9) | 0.271 |
|  | Male |  | 275 (31.4) | 58 (36.0) | 333 (32.1) |  |
| Race | White | 0 | 609 (69.6) | 134 (83.2) | 743 (71.7) | **0.002** |
|  | Black |  | 90 (10.3) | 10 (6.2) | 100 (9.7) |  |
|  | Hispanic |  | 86 (9.8) | 5 (3.1) | 91 (8.8) |  |
|  | Other |  | 90 (10.3) | 12 (7.5) | 102 (9.8) |  |
| BMI | Median (IQR) | 116 | 23.0 (8.0) | 22.0 (5.0) | 23.0 (7.3) | **0.006** |
| ED Admission | Non-ED | 0 | 435 (49.7) | 68 (42.2) | 503 (48.6) | 0.086 |
|  | ED |  | 440 (50.3) | 93 (57.8) | 533 (51.4) |  |
| Discharge Disp. | Home | 0 | 320 (36.6) | 31 (19.3) | 351 (33.9) | **<0.001** |
|  | Psychiatric/Rehab |  | 533 (60.9) | 121 (75.2) | 654 (63.1) |  |
|  | Expired |  | 4 (0.5) | 3 (1.9) | 7 (0.7) |  |
|  | Other |  | 18 (2.1) | 6 (3.7) | 24 (2.3) |  |
| LOS (days) | Median (IQR) | 0 | 1.6 (1.5) | 1.9 (1.7) | 1.6 (1.6) | **<0.001** |
| Insurance Type | Unknown | 0 | 16 (1.8) | 1 (0.6) | 17 (1.6) | 0.681 |
|  | Commercial |  | 781 (89.3) | 145 (90.1) | 926 (89.4) |  |
|  | Government |  | 78 (8.9) | 15 (9.3) | 93 (9.0) |  |
| Suicide Category | Ingestion | 0 | 580 (66.3) | 106 (65.8) | 686 (66.2) | 0.745 |
|  | Hanging/Strangulation |  | 7 (0.8) | 3 (1.9) | 10 (1.0) |  |
|  | Suicidal Ideation |  | 95 (10.9) | 17 (10.6) | 112 (10.8) |  |
|  | Other/Self Harm |  | 28 (3.2) | 4 (2.5) | 32 (3.1) |  |
|  | Self-Harm not Apparent |  | 165 (18.9) | 31 (19.3) | 196 (18.9) |  |
| Hospital Death | Survived | 0 | 871 (99.5) | 158 (98.1) | 1029 (99.3) | 0.080 |
|  | Expired |  | 4 (0.5) | 3 (1.9) | 7 (0.7) |  |
| Death | Survived | 0 | 867 (99.1) | 156 (96.9) | 1023 (98.7) | **0.038** |
|  | Expired |  | 8 (0.9) | 5 (3.1) | 13 (1.3) |  |
| Weekday/<br>Weekend | Weekday | 0 | 662 (75.7) | 124 (77.0) | 786 (75.9) | 0.764 |
|  | Weekend |  | 213 (24.3) | 37 (23.0) | 250 (24.1) |  |
| School Session | School Year | 0 | 674 (77.0) | 112 (69.6) | 786 (75.9) | **0.045** |
|  | Summer Vacation |  | 201 (23.0) | 49 (30.4) | 250 (24.1) |  |
| Time of Day | Night Hours | 0 | 645 (73.7) | 106 (65.8) | 751 (72.5) | **0.044** |
|  | Office Hours |  | 230 (26.3) | 55 (34.2) | 285 (27.5) |  |
| Median Income | Median (IQR) | 56 (%) | 49428.0 | 50232.0 | 49428.0 | 0.097 |
|  |  |  | (18162.0) | (20163.2) | (18160.0) |  |

**Note:** Categorical data is expressed as frequency (percent) and analyzed via Fisher's Exact Test; Numeric data are expressed as median ± interquartile range and analyzed via Mann-Whitney U-Test. P-value of $<0.05$ was considered statistically significant. BMI: basal metabolic index; Discharge Disp: discharge disposition; ED: emergency department; IQR: inter-quartile range; LOS: length of stay; PICU: pediatric intensive care unit.

Temporal variables were computed based on a patient’s hospital admission date and time. Seasonality was qualified as school session: ***either*** *school year or summer vacation*. The months June, July, and August were assumed as general summer vacation months for the study population (School Session = *Summer Vacation*), while the remaining months were considered as school year months (School Session = *School Year*). Furthermore, the day of week of the hospital admission was used to determine whether the admission occurred on a weekend or a weekday; time of admission was defined as: office hours (7:30 a.m. – 5:30 p.m.) versus, night hours (5:30 p.m. – 7:30 a.m.) [11,12].

The patient-level data was supplemented with zip code-aggregated data from the U.S. Census Bureau’s 2017 5-year American Community Survey estimates, and included median household income, various demographic and socioeconomic variables [13].

### 2.3 Statistical Analyses

Quantitative data are expressed as median ± interquartile range. Qualitative data are expressed as frequency (percent). Group comparisons between hospitalized PICU and non-PICU admitted cases for numeric data were implemented using Mann-Whitney U-Tests; qualitative comparisons were made using Fisher’s Exact Tests. *P*-value of <0.05 was considered statistically significant.

Modeling the odds of PICU admission was performed using a binomial logistic regression model [14]. With the large skew between PICU and non-PICU admissions, a multivariable model was selected prioritizing model parsimony to prevent over-fitting and complete model separation.

Zip code tabulation areas (ZCTAs) shapefiles were obtained using the tigris package offered in R statistical software [15]. These shapefiles are pulled from the U.S. Census Bureau’s TIGER database. Frequencies for PICU and non-PICU suicide-related admissions were then calculated for each zip code and standardized based on the U.S. Census Bureau’s 2017 ACS estimates of middle school and high school enrollment to calculate the frequency per 1,000 students [12]. Additional zip-code level information (rural population, percent of family homes, and housing unit estimates) were obtained through the Integrated Public Use Microdata Series (IPUMS) National Historical Geographic Information System [16]. This information was then used to create bivariate choropleth maps in R [17,18], using median values to classify intensity. Relative PICU admissions were calculated for all ZCTAs as the count of standardized PICU admissions divided by the total combined count of standardized PICU and non-PICU admissions and given as a percent (Supplemental Figs 2-4). This allowed for an exploratory approach to examine spatial relationships between the observed zip code suicide-related admissions and aggregate-level demographic information. All analyses were performed in R version 3.6.0 [14].

Given the temporal component of the data (suicide-related admissions over time), basic seasonal decomposition was performed using an equidistant moving average and average values at time points to explore seasonality.

## 3. Results

### 3.1 Demographics

Patients included in the analysis from 1,036, were hospitalized (n=875), and those with a PICU admission (n=161). Initial findings from the demographic analysis revealed statistically significant differences between the patients who were hospitalized versus those admitted to the PICU (Table 1), including: age, race, BMI, discharge disposition, LOS, mortality, school session, and time of day of incident. Age was strongly related to PICU admission; median age of PICU-admitted patients being 16 years and non-PICU-admitted patients having a median age of 15.5 years (p = 0.001) (Figure 1). Although not statistically significant, females consistently appeared more frequently in both PICU and non-PICU admissions (64 %, 68.6 %, respectively). Race/ethnicity yielded significant associations with PICU admissions (p=0.002), as evident in the predominant Caucasian PICU admissions (83 %). Patients admitted to the PICU had lower median BMIs (22.0 vs 23.0, p = 0.006). The suicide category did not strongly associate with PICU admissions (p=0.745). A larger number of non-PICU patients were discharged home after their hospitalization (p = <0.001), spent less time in the hospital (1.6 vs. 1.9; p = <0.001), and were more likely to survive their hospitalization (p = 0.038). In terms of temporality, non-PICU admissions were higher during the school year (p = 0.045) and night hours (p = 0.044).

**Figure 1.**
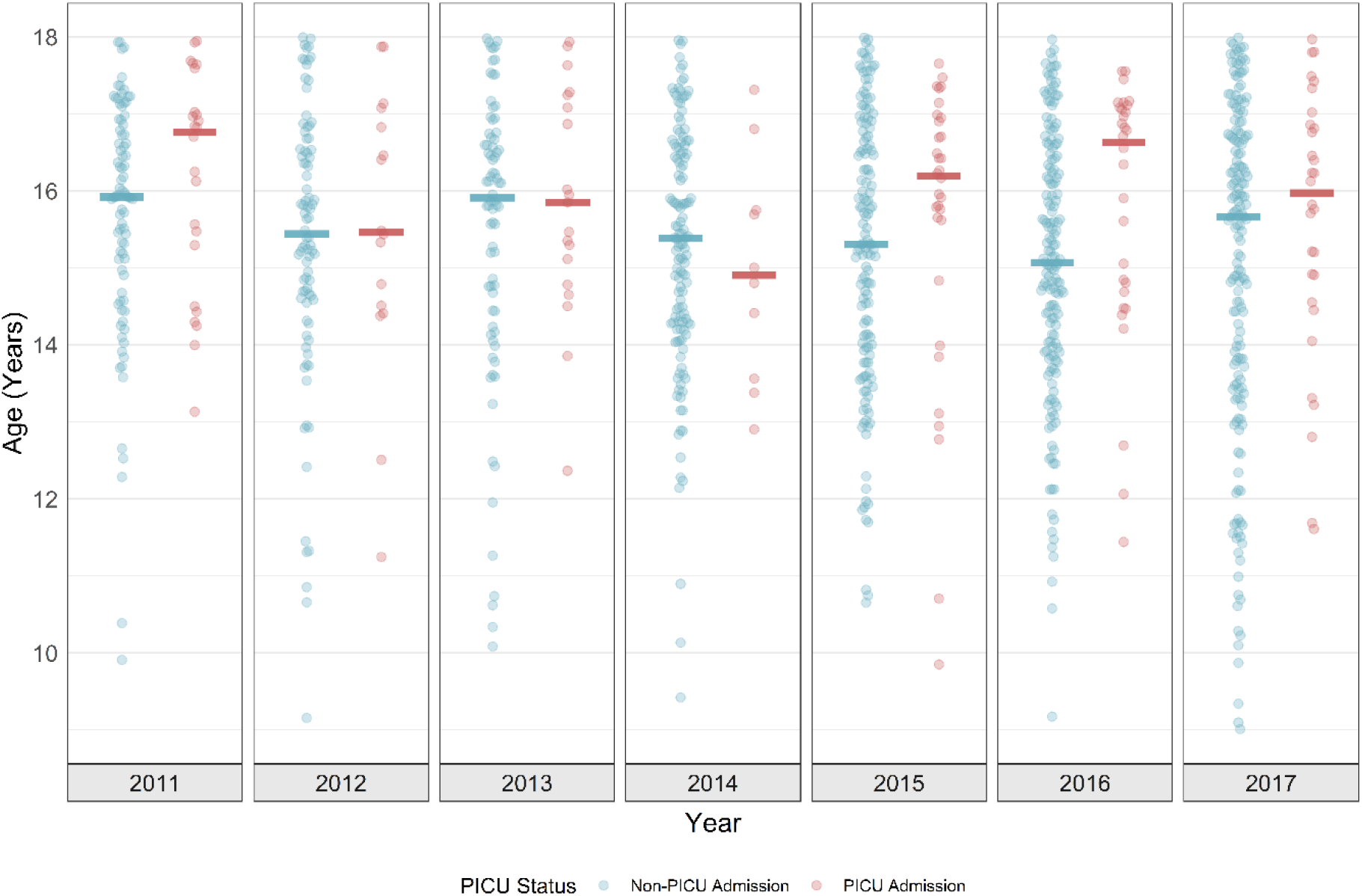
Observed Age Distribution of PICU and Non-PICU Patients (2011-2017)

**Figure 2.**
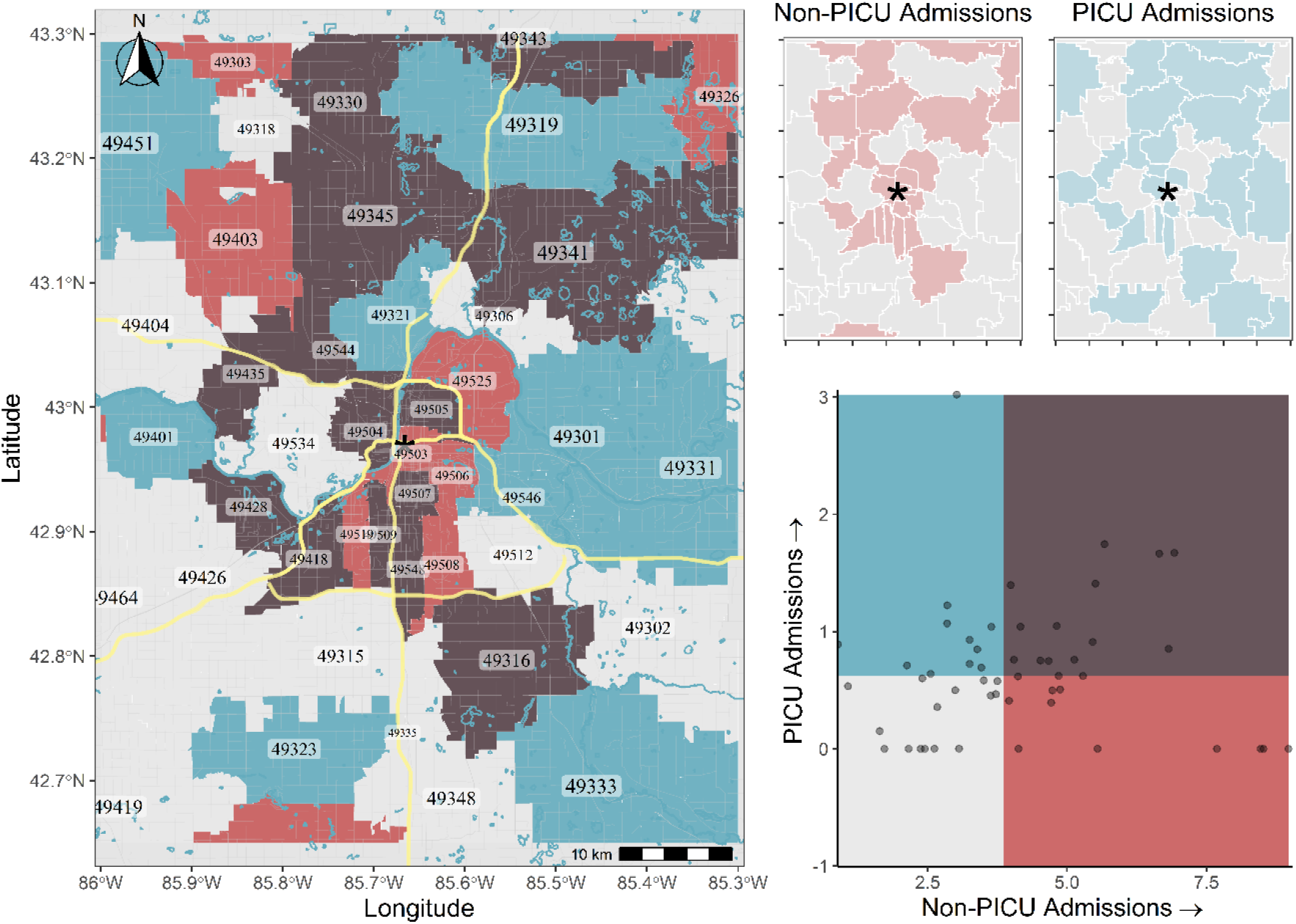
Bivariate Chloropleth Map of PICU and Non-PICU Admissions by 5-Digit Zip Code Standardized per 1,000 Students

### 3.2. Logistic Regression

A binomial logistic regression was run to predict PICU admission while controlling for other covariates. All relevant covariates were assessed in individual univariate models, and a stepwise selection method was used to create a multivariable model while prioritizing model parsimony by minimizing Akaike Information Criterion (AIC) (Table 2).

**Table 2:**
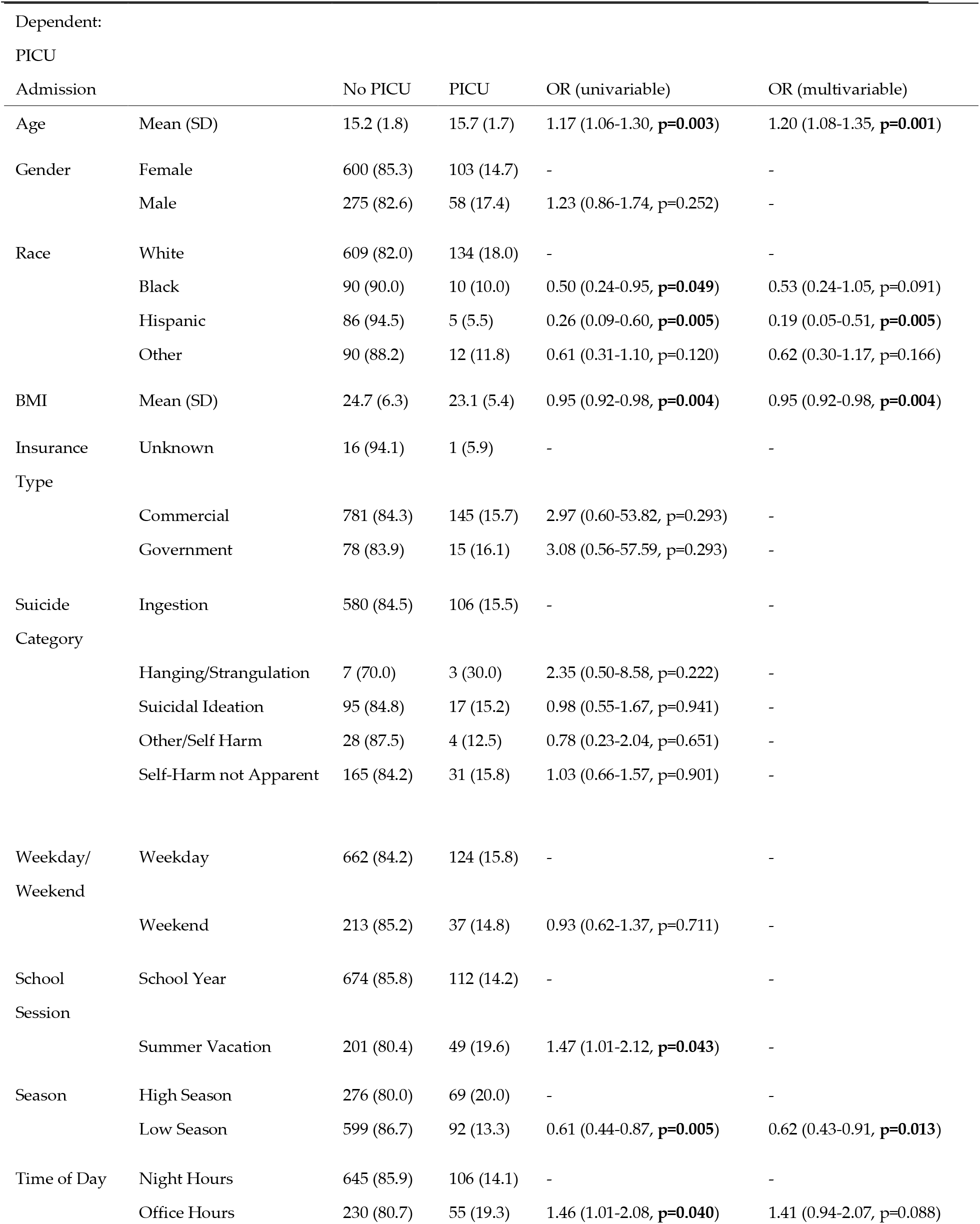
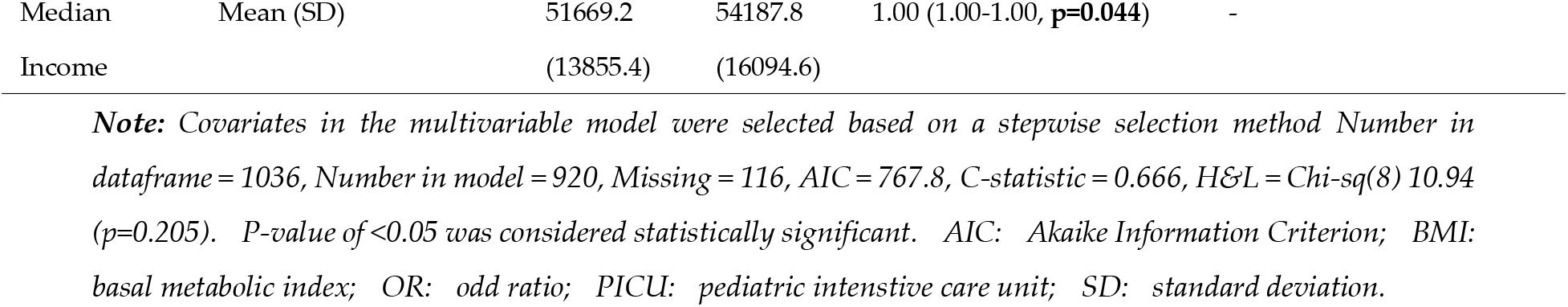
Results of Logistic Regression Analysis for PICU and Non-PICU admissions

| Dependent: |  |  |  |  |  |
| --- | --- | --- | --- | --- | --- |
| PICU |  |  |  |  |  |
| Admission |  | No PICU | PICU | OR (univariable) | OR (multivariable) |
| Age | Mean (SD) | 15.2 (1.8) | 15.7 (1.7) | 1.17 (1.06-1.30, **p=0.003**) | 1.20 (1.08-1.35, **p=0.001**) |
| Gender | Female | 600 (85.3) | 103 (14.7) | - | - |
|  | Male | 275 (82.6) | 58 (17.4) | 1.23 (0.86-1.74, p=0.252) | - |
| Race | White | 609 (82.0) | 134 (18.0) | - | - |
|  | Black | 90 (90.0) | 10 (10.0) | 0.50 (0.24-0.95, **p=0.049**) | 0.53 (0.24-1.05, p=0.091) |
|  | Hispanic | 86 (94.5) | 5 (5.5) | 0.26 (0.09-0.60, **p=0.005**) | 0.19 (0.05-0.51, **p=0.005**) |
|  | Other | 90 (88.2) | 12 (11.8) | 0.61 (0.31-1.10, p=0.120) | 0.62 (0.30-1.17, p=0.166) |
| BMI | Mean (SD) | 24.7 (6.3) | 23.1 (5.4) | 0.95 (0.92-0.98, **p=0.004**) | 0.95 (0.92-0.98, **p=0.004**) |
| Insurance Type | Unknown | 16 (94.1) | 1 (5.9) | - | - |
|  | Commercial | 781 (84.3) | 145 (15.7) | 2.97 (0.60-53.82, p=0.293) | - |
|  | Government | 78 (83.9) | 15 (16.1) | 3.08 (0.56-57.59, p=0.293) | - |
| Suicide Category | Ingestion | 580 (84.5) | 106 (15.5) | - | - |
|  | Hanging/Strangulation | 7 (70.0) | 3 (30.0) | 2.35 (0.50-8.58, p=0.222) | - |
|  | Suicidal Ideation | 95 (84.8) | 17 (15.2) | 0.98 (0.55-1.67, p=0.941) | - |
|  | Other/Self Harm | 28 (87.5) | 4 (12.5) | 0.78 (0.23-2.04, p=0.651) | - |
|  | Self-Harm not Apparent | 165 (84.2) | 31 (15.8) | 1.03 (0.66-1.57, p=0.901) | - |
| Weekday/Weekend | Weekday | 662 (84.2) | 124 (15.8) | - | - |
|  | Weekend | 213 (85.2) | 37 (14.8) | 0.93 (0.62-1.37, p=0.711) | - |
| School Session | School Year | 674 (85.8) | 112 (14.2) | - | - |
|  | Summer Vacation | 201 (80.4) | 49 (19.6) | 1.47 (1.01-2.12, **p=0.043**) | - |
| Season | High Season | 276 (80.0) | 69 (20.0) | - | - |
|  | Low Season | 599 (86.7) | 92 (13.3) | 0.61 (0.44-0.87, **p=0.005**) | 0.62 (0.43-0.91, **p=0.013**) |
| Time of Day | Night Hours | 645 (85.9) | 106 (14.1) | - | - |
|  | Office Hours | 230 (80.7) | 55 (19.3) | 1.46 (1.01-2.08, **p=0.040**) | 1.41 (0.94-2.07, p=0.088) |
| Median | Mean (SD) | 51669.2 | 54187.8 | 1.00 (1.00-1.00, **p=0.044**) | - |
| Income |  | (13855.4) | (16094.6) |  |  |

**Note:** Covariates in the multivariable model were selected based on a stepwise selection method. Number in dataframe = 1036, Number in model = 920, Missing = 116, AIC = 767.8, C-statistic = 0.666, H&L = Chi-sq(8) 10.94 (p=0.205). P-value of <0.05 was considered statistically significant. AIC: Akaike Information Criterion; BMI: basal metabolic index; OR: odd ratio; PICU: pediatric intensive care unit; SD: standard deviation.

The final logit model consisted of race, age, BMI, ED admission, school session, and time of day as covariates predicting PICU admission.

After controlling for all other variables in the model, Black and Hispanic patients were found to have considerably lower odds of a PICU admission when compared to Caucasian patients (OR: 0.49 and 0.17, respectively). This result is in contrast to other findings [19]. Older patients are significantly more likely to be admitted to the PICU (OR: 1.21, p = 0.005), as are patients with smaller body mass indices (OR: 0.95, p = 0.0005). Interestingly, with all other multivariable covariates being held constant, initial admission into ED significantly increases the odds of PICU admission (OR: 1.48, p = 0.039). This relationship is only marginally significant in the univariate analysis. The time of year relative to school session is also significant, with patient encounters occurring during summer vacation (June-August) having significantly higher odds of being admitted into the PICU relative to patient encounters during the school year (OR: 1.51, p = 0.044).

### 3.3 Temporal changes and Time-Series Decomposition

We were interested in determining the incidence of suicide over the years, over the course of seasons, and broken-down into and even lower denominator of time of day. Additive seasonal decomposition was used to assess the trend of relative PICU admission over time (Supplemental Fig 1). Simple decomposition was implemented by using the moving average to isolate the trend of relative PICU admissions, and a seasonal component was calculated by computing an average value of relative PICU admissions for each month across the entire span of the data (2011-2017). Error of the decomposition was then computed by removing the trend and seasonal components from the original time series data.

The trend for the relative PICU admissions fluctuates, with peaks in 2013 followed by a stark drop-off leading into 2014, and then a rise again in mid-2014. The seasonal components show that relative PICU admissions spike in June and August, and then again in December and January (Supplemental Table 2). These increases seem to correspond to periods in which patients are recently released from, or about to resume school after a holiday or summer vacation.

### 3.4 Spatial Relationships & Patterns

Spatial relationships were explored by creating bivariate choropleth maps of zip code geographies. Standardizing for pediatric population size of each zip code tabulation area, a moderately-strong correlation in the spatial intensity of PICU admissions and non-PICU admissions was found (Figure 2). The relative PICU admissions also correlate well with aggregate demographic and economic data from the U.S. Census Bureau, with higher relative PICU admissions correlating with increasing rural population size (Supplemental Fig 2), the percentage-increase in housing units from 2011-2017, and the percentage of family households (Supplemental Fig 4).

## 4. Discussion

Suicidal behavior is determined by many factors, none of which are the immediate focus of medical management when the patient is admitted to the PICU. The priorities are more a determination of risk and providing aggressive measures needed to preserve life and minimize morbidity. Once the patient is stabilized in the PICU setting, patients are transferred to the in-patient floor and returned (directly or after a prolonged hospital stay) to a long-term care facility, or to the previous environment (and set of circumstances) that led to the initial behavior. Studies now show that factors such as school systems, socioeconomic and parental unemployment rates are important in determining suicide ideation, and attempted and completed suicide in children and adolescents [20-22]. In this study, we used the patients in the PICU as the high-risk group and compared them with non-ICU in-patients to extract both clinical and social variables available in the EMR to develop an understanding of SSA that includes community-based risk factors.

Caucasian patients had higher admission rates in our PICU (83.2 %) compared to other race/ethnic groups which is probably a reflection of race demographics in Grand Rapids, MI and the Midwest in general where the Caucasian population is over 82%. Though Caucasians have historically had a higher rate of suicide, there is recent data supporting rising rates of suicide in Black children aged 5-14 [23]. Studies show that Black adolescents have experienced an increase in rates of suicide and lasting injury caused by attempts [5].

In addition, we found BMI to be of significance. Patients in the PICU had significantly lower BMIs than the other patients that were hospitalized for observation. Studies have shown that a higher BMI, though associated with depression, is protective when it comes to SSA [24].

Proxy data in the EMR allowed us to examine the communities of our patients such as zip codes and payer data. This data, when paired with other databases such U.S. Census Bureau, gives us an understanding of potential social drivers for self-directed violence in our patients.

There are 248 cities in the US with a population of 100-300,000 people making up an eighth of the entire population and this list includes some of the fastest growing cities [25]. Grand Rapids, with a population of 201,000, is one of these cities. We examined the areas around Grand Rapids and found that there were hotspots that could clearly be identified based on PICU suicide-related admissions. The hotspots were outside the city limits and included regions that are in rapid growth and transition. As the city becomes more urbanized, the regions on the outskirts are subject to unique stressors that need better characterizing. The areas around Grand Rapids are experiencing some of the greatest growths in population in Michigan. Kent County, which is the County Seat (governmental center) of Grand Rapids, has increased in population at rates over 9% from 2010 to 2019 [26]. Population increases will change living conditions not only in the cities themselves, but adjoining neighborhoods. Our geospatial examination has shown that suicide increases rapidly in rural and suburban areas compared to urban ones (Supplemental Fig 2). This could simply be a product of a denser pediatric population in urban regions leading to a higher denominator, and thus lower rates, or reflect a scarcity of mental health resources in these areas [3]. Additional steps need to be taken to better understand the geospatial factors around pediatric suicide.

Five-digit zip code-tabulation of rural/suburban areas includes large spatial heterogeneity of populations. Future work could entail assessing smaller geographic areas, such as census block-groups to gain a more granular understanding around the immediate environment from which these suicide events emerge. Meyer, et al (2016) used point-pattern modelling with aggregate demographic information obtained from small, administrative quarters in Zurich, CH in Europe, to better understand factors associated with psychiatric hospital admissions [27]. This approach could easily be adapted for assessing geographic patterns of suicide-related hospital admissions.

Evidence suggests that schools influence children and adolescents health behaviors for self-harm and suicide [28]. However, data exists in the EMR that allows us to examine the influence of the school year on our patients lives. We did this by applying basic time-series decomposition and found that increases in rates of PICU admissions seem to crest when schools are in recess or just about to start. We know that the nature and availability of social ties which are primarily gained from school early on, have impact on the higher functioning of adolescents. Targeted programs or interventions that originate from schools may alleviate some of the stress these patients feel at transition either from recess or getting back to school. Connecting actual suicide rates in the geographical hotspots to actual school systems is beyond the scope of this paper, as is any commentary on recommended interventions.

Developing an understanding of the communities is important for another reason. The most robust risk factor associated with suicide death is a previous suicide attempt [5]. A suicide attempt is a positive predictor for a completed suicide [29]. Then it becomes important to assess the communities and the support available to this child before returning him or her to that community. There may be a need to review how we think and screen for suicide in terms of age and practice, as it has been found that 45 % of suicide patients were seen in a clinical setting in the month prior to their death, versus less than half that number (19 %) who were in contact with their mental health providers [30].

If high risk regions are identified, then active screening by physicians and advanced practitioners in those communities may help alleviate some of the hospital admissions. In a non-ambulatory setting, the Columbia-Suicide Severity Rating Scale (C-SSRS) is the recommended method for suicide screening and risk stratification [31]. These recommendations align with the Joint Commissions National Patient Safety Goal (NPSG.15.01.01) to reduce the risk for suicide [32]. Substantially more people are hospitalized as a result of nonfatal suicidal behavior than are fatally injured, with a substantial number treated in an ambulatory setting [33]. These patients are considered an *at-risk* group but could benefit from being treated as a *high-risk* group to increase the likelihood of receiving early intervention with mental health services [34]. The recommendation for suicide screening practices for patients 10 and older in Western Michigan according to the Pediatric Behavioral Health Screening Practice Excellence Workgroup of HDVCH includes: 1) in the ambulatory setting, Q9 of the Patient Health Questionnaire-9 (PHQ-9) [35,36] 2) the Columbia Suicide Severity Rating Scale (C-SSRS) with further assessment and triage stratification; any answer other than “none” is considered a positive screen on Q9 of the PHQ-9.

One limitation of the work is the nature of the experimental design. Future work may include a prospectively collected dataset, to avoid any selection bias in the patient population.

## 5. Conclusions

Social determinants of suicide are likely to contribute as much as, if not more than, individual risk factors, but they have been poorly studied to date. This study highlights geospatial and temporal analysis as a means of exploring community risk to develop a better understanding to assist us in providing for the mental health care needs of our communities for future interventions.

## Data Availability

Data is available upon request.

## Supplementary Materials

Figure S1: Trend of relative PICU admission over time; Table S1: Data Dictionary; Figure S2: Higher Relative PICU Admissions Correlating with Increases Rural Population Size; Table S2: Median relative frequency of PICU admissions for each month, accompanied by seasonal decomposition; Figure S3: Standardized PICU and non-PICU Admissions Relative to Housing; Figure S4: Standardized PICU and non-PICU Admissions Relative to Family Households.

## Author Contributions

Conceptualization, S.R. and M.L-L.; methodology, A.K., S.R. and M.L-L.; software, A.K.; validation, A.K.; formal analysis, S.R., A.K.; investigation, S.R., A.K., A.B., M.L-L.; resources, B.E.; data curation, M.O., A.H., A.M.E.V.; writing—original draft preparation, A.K, M.O., A.M.E.V., B.E., A.H., A.B., M.L-L., S.R.; writing—review and editing, A.K, M.O., A.M.E.V., B.E., A.H., A.B., M.L-L., S.R.; visualization, A.K., S.R.; supervision, S.R., M.L-L.; project administration, B.E., M.L-L.; funding acquisition, N/A. All authors have read and agreed to the published version of the manuscript.

## Acknowledgments

The authors would like to thank the PICU staff at Helen DeVos Children’s Hospital for their support in the completion of this study and various contributions, and the Pediatric Behavioral Health Screening Practice Excellence Workgroup: Kiran Taylor MD, Division Chief, Psychiatry and Behavioral Medicine (Sponsor), Jennifer Bowden, MD, Hannah Jaworski, MD, Lisa Lowery, MD, Phillip Waalkes, DO, Jody Sprague, LMSW, Erica Auger, LMSW, Scott Stebbins, LMSW, Sarah Pentoney LMSW, Ashleigh Nurski, MSN, Ian Straayer, Laura Stuursma, MBA, Drew Peklo, MSN, Christina Pries, DNP, Stephanie Mullennix, MSN, and Beth Kuik, LMSW. The authors would also like to thank Jessica Parker, MS, for her contributions in initiating the data acquisition and statistical summaries.

## References

1. Crosby, A.E., Ortega, L., Melanson, C. Self-directed Violence Surveillance: Uniform Definitions and Recommended Data Elements, Version 1.0. Control, N.C.f.I.P.a., Ed. Centers for Disease Control and Prevention: Atlanta (GA), 2011.

2. Gvion, Y., Levi-Belz, Y. Serious Suicide Attempts: Systematic Review of Psychological Risk Factors. Front Psychiatry 2018, 9, 56, doi:10.3389/fpsyt.2018.00056.

3. Kegler, S.R., Stone, D.M., Holland, K.M. Trends in Suicide by Level of Urbanization - United States, 1999-2015. MMWR Morb Mortal Wkly Rep 2017, 66, 270–273, doi:10.15585/mmwr.mm6610a2.

4. Patton, G.C., Coffey, C., Sawyer, S.M., Viner, R.M., Haller, D.M., Bose, K., Vos, T., Ferguson, J., Mathers, C.D. Global patterns of mortality in young people: a systematic analysis of population health data. Lancet 2009, 374, 881–892, doi:10.1016/S0140-6736(09)60741-8.

5. Curtin, S.C., Warner, M., Hedegaard, H. Increase in Suicide in the United States, 1999-2014. NCHS Data Brief 2016, 1–8.

6. Fontanella, C.A., Hiance-Steelesmith, D.L., Phillips, G.S., Bridge, J.A., Lester, N., Sweeney, H.A., Campo, J.V. Widening rural-urban disparities in youth suicides, United States, 1996-2010. JAMA pediatrics 2015, 169, 466–473, doi:10.1001/jamapediatrics.2014.3561.

7. Joe, S., Banks, A., Belue, R. Suicide risk among urban children. Children and Youth Services Review 2016, 68, 73-79, doi:https://doi.org/10.1016/j.childyouth.2016.07.002.

8. Woo, J.-M., Okusaga, O., Postolache, T.T. Seasonality of suicidal behavior. Int J Environ Res Public Health 2012, 9, 531–547, doi:10.3390/ijerph9020531.

9. Pollack, M.M., Patel, K.M., Ruttimann, U.E. PRISM III: an updated Pediatric Risk of Mortality score. Crit Care Med 1996, 24, 743–752.

10. Slater, A., Shann, F., Pearson, G., Paediatric Index of Mortality Study, G. PIM2: a revised version of the Paediatric Index of Mortality. Intensive Care Med 2003, 29, 278–285, doi:10.1007/s00134-002-1601-2.

11. Venables, W.N., Ripley, B.D. Modern Applied Statistics with S, 4th Edition ed., Springer: New York, 2002.

12. Bureau, U.S.C. American Community Survey, 2016 American Community Survey 5-Year Estimates, Table DP02. Andy Kampfschulte: HDVCH, 2019; p using American FactFinder.

13. Harrison, E., Drake, T., Ots, R. finalfit: Quickly Create Elegant Regression Tables and Plots when Modelling R, 2019.

14. Team, R.C. R: A language and environment for statistical computing, Vienna, Austria, 2018.

15. Walker, K. tigris: Load Census TIGER/Line Shapefiles, 2018.

16. Manson, S., Schroeder, J., Van Riper, D., Ruggles, S. IPUMS National Historical Geographic Information System: Version 14.0. Minneapolis, MN: IPUMS., 2019; http://doi.org/10.18128/D050.V14.0.

17. Prener C; Grossenbacher, T., Zehr, A., Stevens, J. Tools and Palettes for Bivariate Thematic Mapping, 2020.

18. Wickham, H. ggplot2: Elegant Graphics for Data Analysis, Springer: Verlag, New York, 2016.

19. Lindsey, M.A., Sheftall, A.H., Xiao, Y., Joe, S. Trends of Suicidal Behaviors Among High School Students in the United States: 1991-2017. Pediatrics 2019, 144, doi:10.1542/peds.2019-1187.

20. Fang, M. School poverty and the risk of attempted suicide among adolescents. Social psychiatry and psychiatric epidemiology 2018, 53, 955–967, doi:10.1007/s00127-018-1544-8.

21. Benbenishty, R., Astor, R.A., Roziner, I. A School-Based Multilevel Study of Adolescent Suicide Ideation in California High Schools. J Pediatr 2018, 196, 251–257, doi:10.1016/j.jpeds.2017.12.070.

22. Aleck, O., Stefania, M., James, T., James, D., Ruth, H., Lisa, C., Amber, L., Clyde, H. The impact of fathers’ physical and psychosocial work conditions on attempted and completed suicide among their children. BMC public health 2006, 6, 77, doi:10.1186/1471-2458-6-77.

23. Bridge, J.A., Horowitz, L.M., Fontanella, C.A., Sheftall, A.H., Greenhouse, J., Kelleher, K.J., Campo, J.V. Age-Related Racial Disparity in Suicide Rates Among US Youths From 2001 Through 2015. JAMA Pediatr 2018, 172, 697–699, doi:10.1001/jamapediatrics.2018.0399.

24. Bjørngaard, J.H., Carslake, D., Lund Nilsen, T.I., Linthorst, A.C., Davey Smith, G., Gunnell, D., Romundstad, P.R. Association of Body Mass Index with Depression, Anxiety and Suicide-An Instrumental Variable Analysis of the HUNT Study. PLoS One 2015, 10, e0131708, doi:10.1371/journal.pone.0131708.

25. Resident Population Estimates for the 100 Fastest-Growing U.S. Counties with 10,000 or More Population in 2010: April 1, 2010 to July 1, 2019 (CO-EST2019-CUMGR) U.S. Census Bureau.: 2020; https://www.census.gov/data/tables/time-series/demo/popest/2010s-counties-total.html.

26. Cumulative Estimates of Resident Population Change and Rankings for Counties in Michigan: April 1, 2010 to July 1, 2019 (CO-EST2019-CUMCHG-26). U.S. Census Bureau.: 2020; https://www.census.gov/data/tables/time-series/demo/popest/2010s-counties-total.html.

27. Meyer, S., Warnke, I., Rössler, W., Held, L. Model-based testing for space–time interaction using point processes: An application to psychiatric hospital admissions in an urban area. Spatial and Spatio-temporal Epidemiology 2016, 17, 15-25, doi:https://doi.org/10.1016/j.sste.2016.03.002.

28. Evans, R., Hurrell, C. The role of schools in children and young people’s self-harm and suicide: systematic review and meta-ethnography of qualitative research. BMC public health 2016, 16, 401, doi:10.1186/s12889-016-3065-2.

29. Bostwick, J.M., Pabbati, C., Geske, J.R., McKean, A.J. Suicide Attempt as a Risk Factor for Completed Suicide: Even More Lethal Than We Knew. Am J Psychiatry 2016, 173, 1094–1100, doi:10.1176/appi.ajp.2016.15070854.

30. Luoma, J.B., Martin, C.E., Pearson, J.L. Contact with mental health and primary care providers before suicide: a review of the evidence. Am J Psychiatry 2002, 159, 909–916, doi:10.1176/appi.ajp.159.6.909.

31. Posner, K., Brown, G.K., Stanley, B., Brent, D.A., Yershova, K.V., Oquendo, M.A., Currier, G.W., Melvin, G.A., Greenhill, L., Shen, S., et al. The Columbia-Suicide Severity Rating Scale: initial validity and internal consistency findings from three multisite studies with adolescents and adults. Am J Psychiatry 2011, 168, 1266–1277, doi:10.1176/appi.ajp.2011.10111704.

32. Commission, T.J. R3 Report Issue 18: National Patient Safety Goal (NPSG) 15.01.01 for suicide prevention. 2019; https://www.sprc.org/resources-programs/r3-report-issue-18-national-patient-safety-goal-suicide-prevention#:~:text=R3%20Report%20Issue%2018%3A%20National%20Patient%20Safety%20Goal,who%20are%20identified%20as%20high%20risk%20for%20suicide.

33. Crosby, A.E., Han, B., Ortega, L.A., Parks, S.E., Gfroerer, J., Centers for Disease, C., Prevention. Suicidal thoughts and behaviors among adults aged >/=18 years--United States, 2008-2009. MMWR Surveill Summ 2011, 60, 1–22.

34. Sheftall, A.H., Asti, L., Horowitz, L.M., Felts, A., Fontanella, C.A., Campo, J.V., Bridge, J.A. Suicide in Elementary School-Aged Children and Early Adolescents. Pediatrics 2016, 138, doi:10.1542/peds.2016-0436.

35. Na, P.J., Yaramala, S.R., Kim, J.A., Kim, H., Goes, F.S., Zandi, P.P., Vande Voort, J.L., Sutor, B., Croarkin, P., Bobo, W.V. The PHQ-9 Item 9 based screening for suicide risk: a validation study of the Patient Health Questionnaire (PHQ)-9 Item 9 with the Columbia Suicide Severity Rating Scale (C-SSRS). J Affect Disord 2018, 232, 34–40, doi:10.1016/j.jad.2018.02.045.

36. Gilbody, S., Richards, D., Brealey, S., Hewitt, C. Screening for depression in medical settings with the Patient Health Questionnaire (PHQ): a diagnostic meta-analysis. J Gen Intern Med 2007, 22, 1596–1602, doi:10.1007/s11606-007-0333-y.

