## Supplemental for "Understanding Suicide in Our Community Through the Lens of the Pediatric ICU: An Epidemiological Review (2011-2017) of One Midwestern City in the US"

Article

**Supplementary Materials:**

**Figure S1**: Trend of relative PICU admission over time

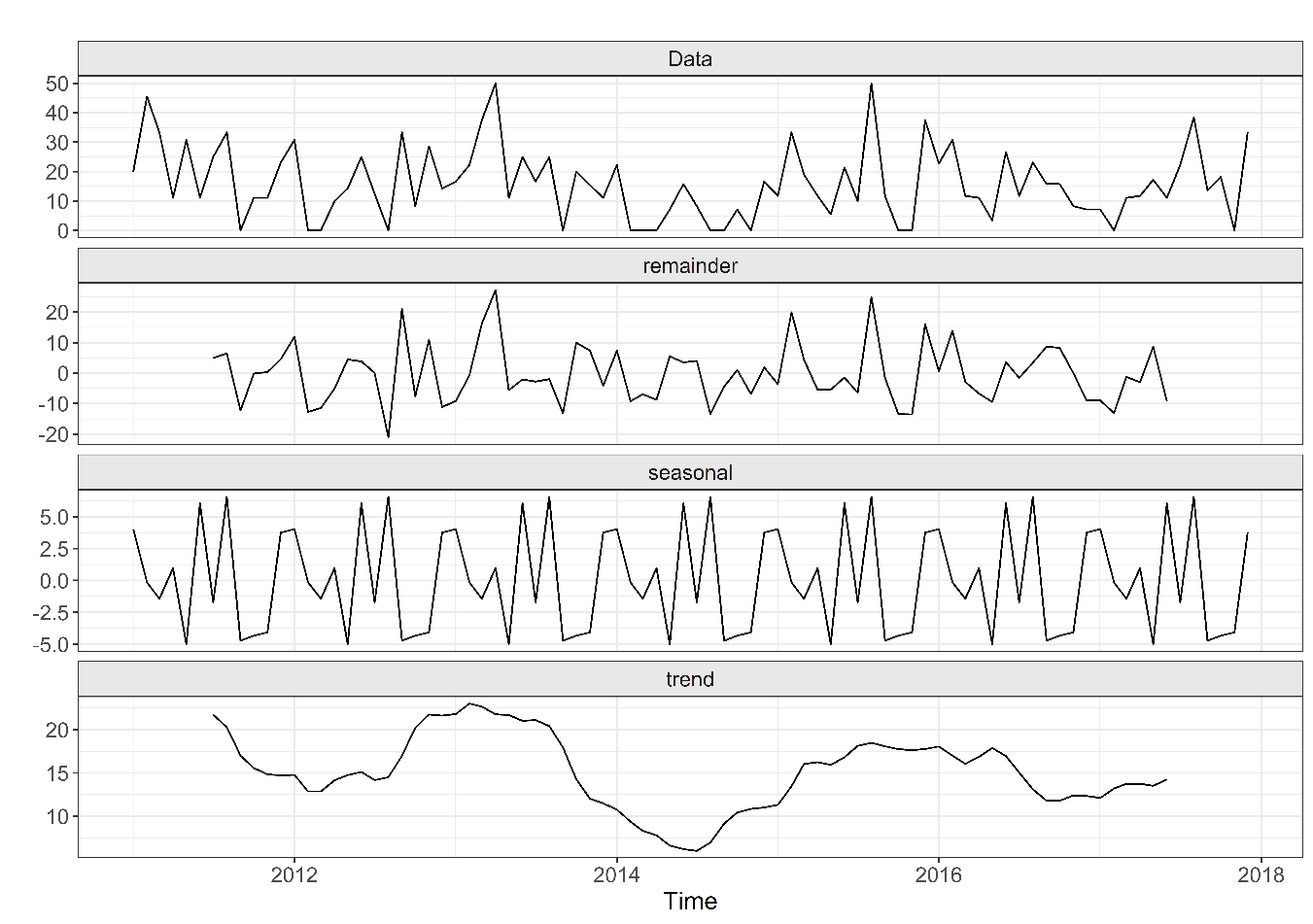

**Table S1:** Data Dictionary

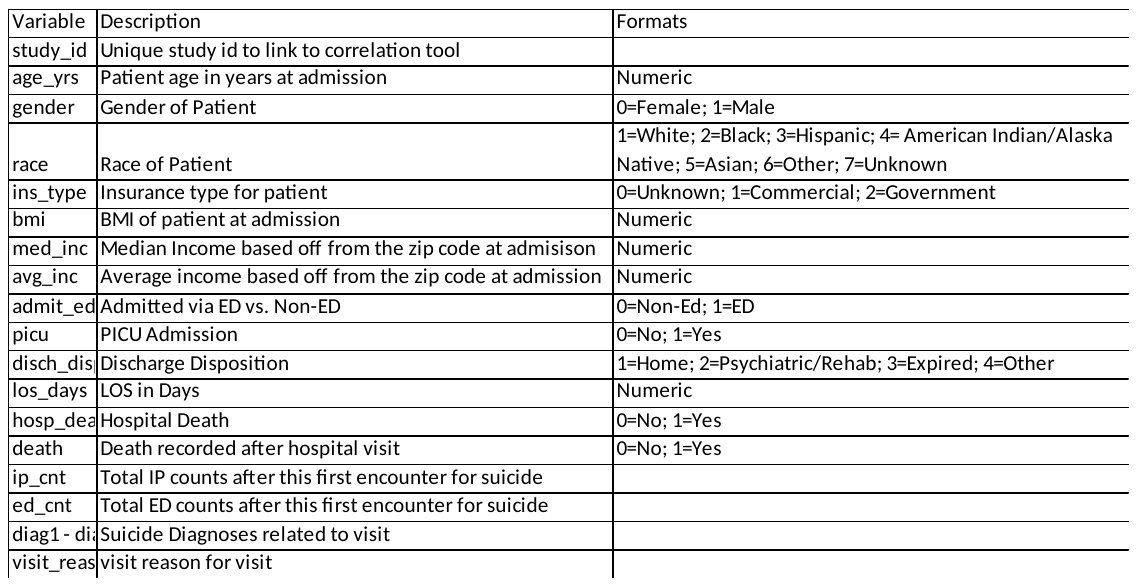

**Figure S2:** Higher Relative PICU Admissions Correlating with Increases Rural Population Size

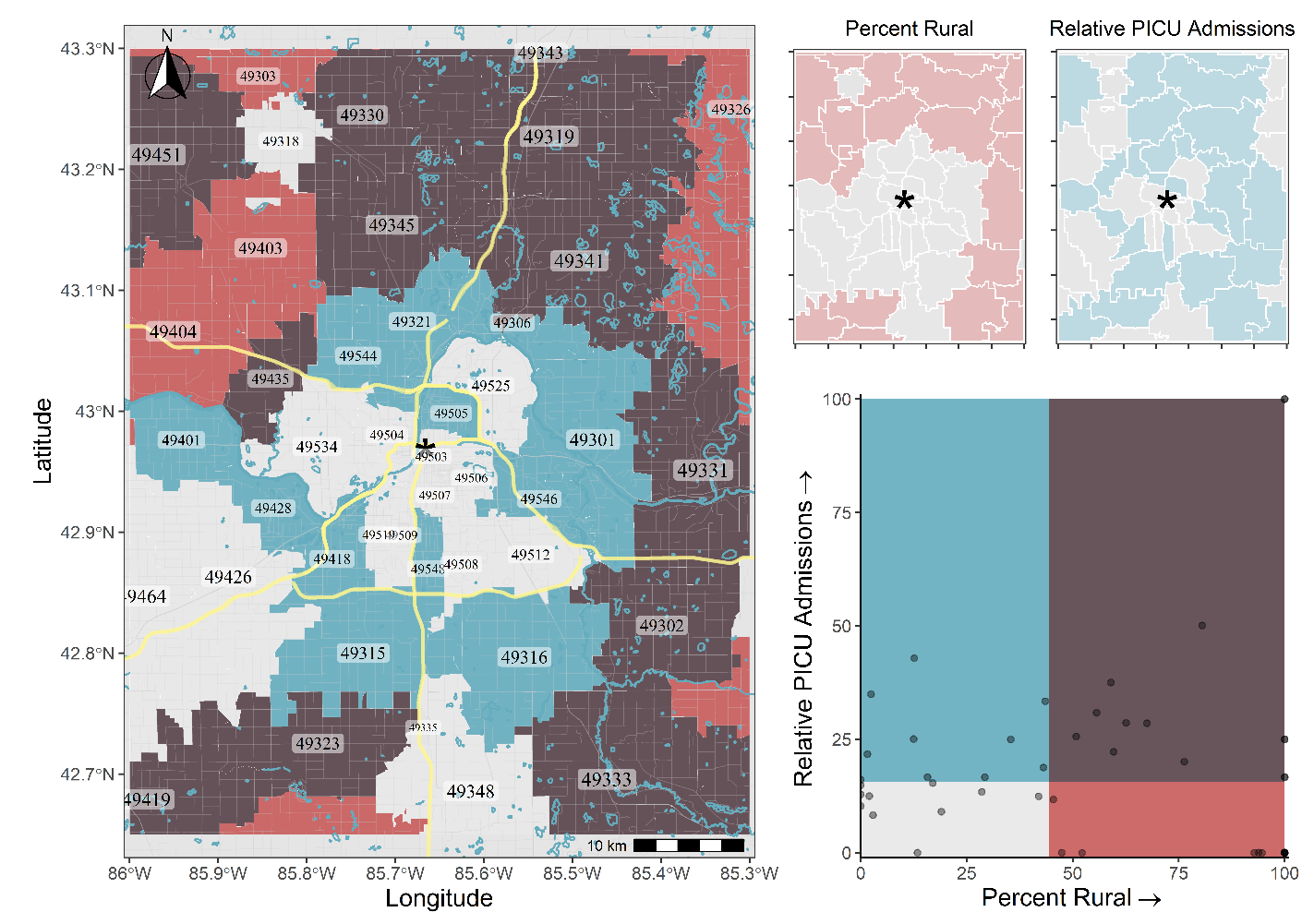

**Table S2:** Median relative frequency of PICU admissions for each month, accompanied by seasonal decomposition.

| **Month** | **Median Relative**  **Frequency** | **Seasonal**  **Adjustment** |
| --- | --- | --- |
| January | 20.00 | 4.03 |
| February | 22.22 | -0.15 |
| March | 11.76 | -1.43 |
| April | 11.11 | 0.96 |
| May | 11.11 | -4.98 |
| June | 21.43 | 6.06 |
| July | 12.50 | -1.70 |
| August | 25.00 | 6.56 |
| September | 11.76 | -4.72 |
| October | 11.11 | -4.32 |
| November | 8.33 | -4.06 |
| December | 16.67 | 3.76 |

**Figure S3:** Standardized PICU and non-PICU Admissions Relative to Housing.

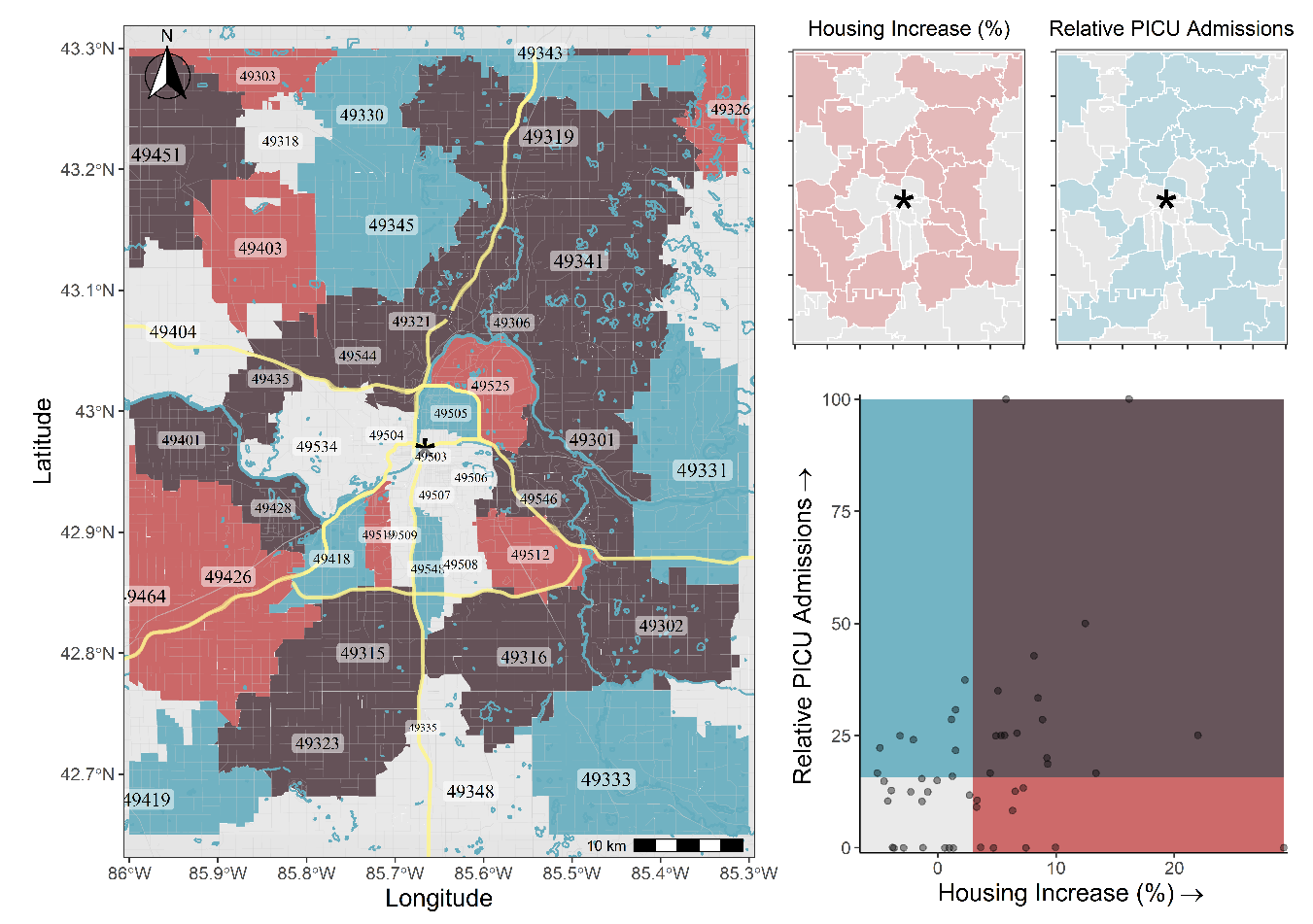

*Note: to examine spatial relationships between the observed zip code suicide-related admissions and aggregate-level demographic information.*

**Figure S4:** Standardized PICU and non-PICU Admissions Relative to Family Households.

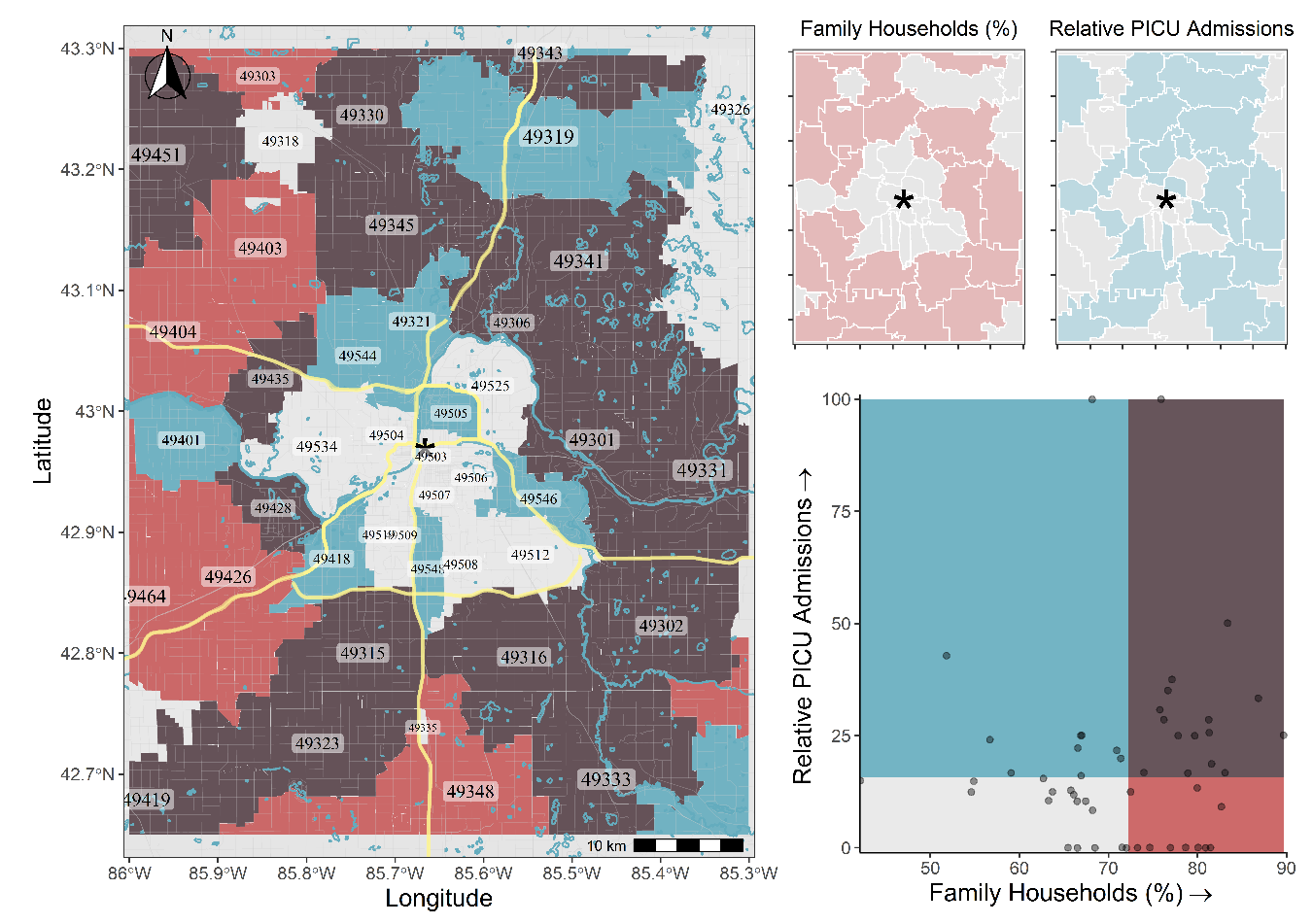
